# Emergency Care in India: A Retrospective Cross-sectional Analysis of Health Management and Information System and Global Burden of Disease

**DOI:** 10.1101/2024.08.16.24312130

**Authors:** Gaurav Urs, Siddhesh Zadey, Padmavathy Krishna Kumar, Anoushka Arora, Tejali Gangane, Pushkar Nimkar, Catherine Staton, Joao Ricardo Nickenig Vissoci

## Abstract

**Background:** To understand the burden of emergency medical conditions (EMCs) and healthcare service utilization, we assessed Emergency Medicine Department (EMD) data from India’s Health Management Information System (HMIS) and EMCs from the Global Burden of Disease (GBD).

**Methods:** This was a retrospective cross-sectional analysis of HMIS and GBD data for 2019. We extracted EMD registrations, hospital admissions, and deaths from HMIS and incidence and deaths from GBD for 29 EMCs at the national and state levels. We analyzed HMIS and GBD data to estimate proportions and rates of registrations, deaths, and incident cases relative to population counts and hospital admissions.

**Results:** In 2019, 119,103,358 patients (8,935.66 per 100,000 people) were registered at EMDs per HMIS. The national EMD registration rate was 6,744.22 per 100,000 hospital admissions, and the EMD death rate was 30,526.36 per 100,000 hospital deaths. Only 12.14% of all HMIS registrations had cause-specific data. GBD estimates that EMCs accounted for 27.22% of all-cause incidence and 51.72% of all-cause mortality. Trauma-related registrations were 8.22% in HMIS, while injuries in GBD accounted for 7% of EMCs. Overall, HMIS EMD registrations were lower than GBD EMC incidence numbers, with regional variations.

**Conclusion:** The study reveals gaps between EMD utilization and EMC disease burden estimates. Improved data integration and reporting can address regional disparities.

## INTRODUCTION

Globally, 90% of deaths related to Emergency Medical Conditions (EMCs) could have been prevented with timely emergency care, highlighting the crucial role of Emergency Medicine Departments (EMDs).^1^ India ranks 144^th^ among 195 countries in terms of access to and capacity for immediate care during medical emergencies, as assessed by the Global Burden of Disease (GBD).^1,2^ India’s pre-hospital and hospital-level emergency department services are fragmented at the state and district levels due to variations in patient volumes, emergency care protocol implementation, and availability of resources such as ambulances or blood banks.^3^ While research can help strengthen emergency care delivery in India and other low- and middle-income countries (LMICs), studies have revealed challenges such as inadequate data collection, ethical research practices, and insufficient funding for research.^4,5^ In India, there are no standard database records of population-level outcomes on EMC disease burden at the state and national levels. The Indian Health Management Information System (HMIS) datasets collect data from health facilities and aggregate counts to provide totals for emergency medical department (EMD) registrations, including classifications for trauma, burns, snake bites, acute cardiac events, cerebrovascular accidents, and obstetric complications.^6^

The GBD 2019 was a large-scale research initiative led by the Institute for Health Metrics and Evaluation (IHME) that estimated epidemiological parameters for 300+ diseases, injuries, and risk factors for 204 countries and territories using advanced statistical modeling methods.^7^ HMIS and GBD serve distinct roles within the health information framework. HMIS aims to record localized data from individual health facilities. In contrast, GBD provides a population-level perspective on disease burden by combining real-world data with statistical modeling to estimate health outcomes across diverse populations and regions. To understand the EMC disease burden and EMD service utilization, we conducted a retrospective cross-sectional analysis of the Indian emergency care system using both HMIS and GBD datasets. We had three objectives: Firstly, we evaluated EMD registrations, deaths, and cause-specific registrations, as measured by HMIS, for total admissions and population rates during 2019-2020. Secondly, we evaluated the incidence and mortality for 29 EMCs modeled in GBD 2019. We then analyzed specific outcome metrics in HMIS and GBD, including EMD service utilization and EMC disease burden, which were expressed as counts, rates, and proportions for population- and cause-specific analyses. Lastly, we assessed data consistency and organization between HMIS and GBD and analyzed similarities and differences in metrics derived from each source.

## METHODS

### Data sources and extraction

The Indian Health Management Information System (HMIS) collects data from health facilities that report monthly on various primary, secondary, and tertiary indicators, including patient admissions, diagnoses, treatments, and outcomes, and consolidates them at the district, state, and national levels. We extracted data on EM registrations, inpatient admissions in EMD, and deaths, as well as all-cause inpatient admissions, outpatient admissions, and hospital deaths for 2019-20. For rates, we used the state-wise mid-year projections for 2019, based on rural, urban, and total populations from the 2011 Census of India.^8^

The GBD uses data from various sources, including vital registration records, national health surveys, health information systems, hospital records, death certificates, and disease registries, to cover a range of health conditions and demographic groups. GBD studies use advanced statistical models to impute and correct missing data and to compensate for variations in reporting quality, thereby helping mitigate bias.^7^

We used the framework by Chang and colleagues that defined EMCs as “diseases or injuries that, if not promptly diagnosed and treated, typically within hours to days, can result in significant physical or mental disability or even death.” This framework was designed to systematically quantify the global burden of emergency care by reviewing all GBD categories against this definition, identifying 31 EMCs.^1^ We extracted data on these 31 EMCs, which closely align with conditions generally managed in EMDs. Since India reported no cases of yellow fever or Ebola in 2019, we were left with 29 EMCs.

We extracted counts and rates (per 100,000 people) for cause-specific and all-cause incidence and deaths, and for the 29 EMCs from GBD 2019.

### Outcomes and Analysis

To assess EMD service utilization, we used HMIS data to develop national- and state-level metrics, including case registration rates and death rates recorded at EMDs. We calculated population-level registration case rates and death rates (per 100,000 people) using mid-year population projections from the census. Because India lacks a population-level, dedicated EMC database, we used GBD-estimated incidence and mortality rates at the state and national levels to assess the EMC disease burden in India.

**Supplementary Table 1** lists EMC outcomes calculated from the GBD and EMD outcomes from HMIS, along with their definitions and formulae.

We mapped overlapping conditions between GBD and HMIS to analyze cause-wise EMC incidence and EMD registration rates (**Supplementary Table 2**). Next, we analyzed case burden using HMIS EMD registration proportions and GBD EMC incidence proportions. Finally, we assessed the mortality burden using death rates from the HMIS EMD and GBD EMC databases. No ethics approval was needed as we used publicly available aggregate-level datasets such as HMIS and GBD.

## RESULTS

### HMIS Descriptive Analysis

From April 2019 to March 2020, 119,103,358 patients (8,935.66 per 100,000 people) were registered at EMDs, according to the Indian HMIS, for a population of 13,32,900,000. Public facilities recorded 114,180,292 (95.87%) of these registrations.

### HMIS EMD Registration Rate

The national HMIS EMD registration rate was 6,744.22 registrations per 100,000 hospital admissions, with Manipur having the highest rate at 30,124.86 and Rajasthan the lowest at 1,210.76. Manipur, Arunachal Pradesh, and Mizoram in the Northeast had higher registration rates than North Indian states such as Rajasthan and West Indian states such as Gujarat. Goa, a small West Indian state in both geography and population, however, stood out for its high registration rate. Meghalaya and Nagaland had higher registration rates than their neighbors. Kerala, Karnataka, Tamil Nadu, Telangana, and Andhra Pradesh had moderate to high registration rates, with Kerala leading the way (**Figure 1A**).

**Figure 1:**
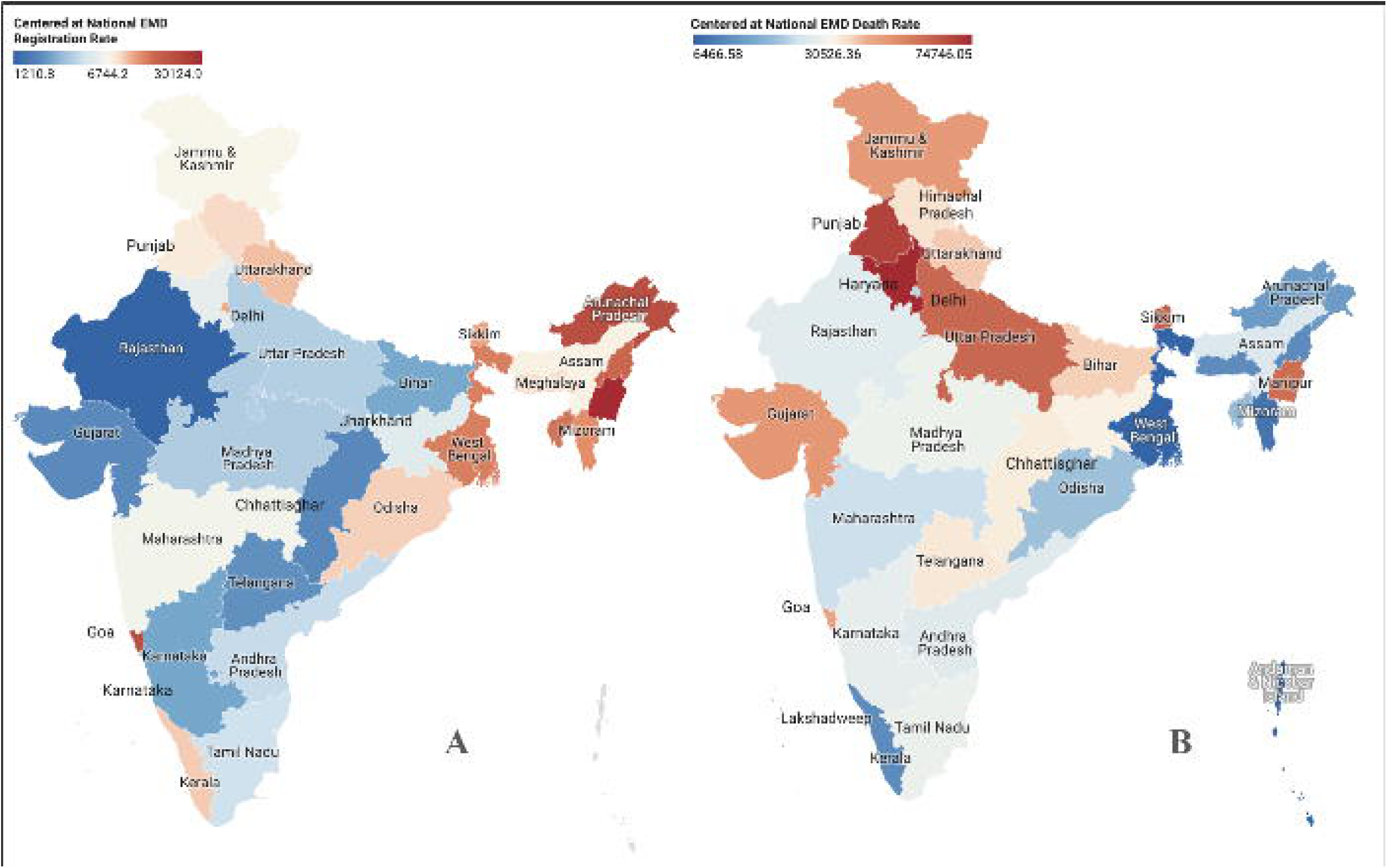
HMIS Emergency Medicine Department Rates for 35 States and Union Territories (2019-20), including A) Registration Rates and B) Death Rates.

### HMIS EMD Death Rate

The national HMIS EMD death rate was 30,526.36 deaths per 100,000 hospital deaths, with Haryana having the highest rate at 74,746.05 and West Bengal having the lowest at 6,466.56. Mizoram, Arunachal Pradesh, and Nagaland in the Northeast had lower death rates than Haryana and Punjab in North India, as well as Kerala and Karnataka in the South. Goa, like before, stood out for its relatively high death rate (**Figure 1B**).

### HMIS Number of Registration Cause-Wise and Cause-Specific Registration Proportion

Among HMIS EMD registrations nationwide, only 12.14% had category-specific data. Trauma accounted for 9,789,272 (8.22%) EMD registrations, followed by obstetric complications (2,221,216 or 1.86%), acute cardiac events (968,374 or 0.81%), cerebrovascular accidents (670,878 or 0.56%), snake bites (518,674 or 0.44%), and burns (287,088 or 0.24%) (**Figure 2A**).

**Figure 2:**
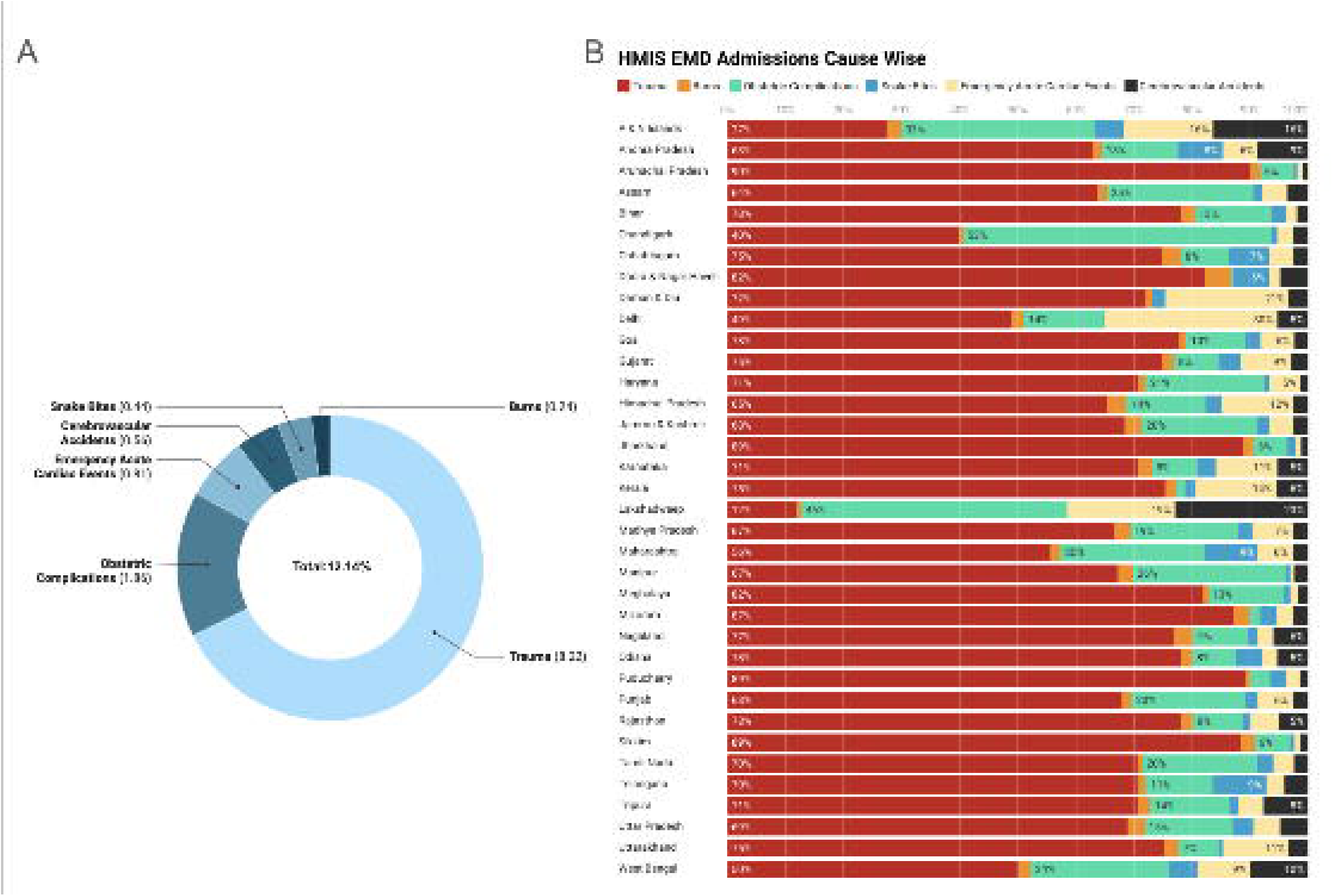
HMIS Emergency Medicine Department Cause-Specific Registration Numbers (2019­20) at the A) National and B) State levels.

Within states, EMD registrations varied significantly by cause. Arunachal Pradesh (90%), Puducherry (89.1%), Jharkhand (89%), Sikkim (89%), and Mizoram (87%) had the highest percentages of trauma-related registrations. Chandigarh had the highest percentage of obstetric cases (53%), followed by Lakshadweep (46%), Andaman and Nicobar Islands (33%), and Manipur (26%). Dadra and Nagar Haveli (4.3%) had the highest number of burn cases registered, followed by Himachal Pradesh (3.6%), Nagaland (3.5%) and Chhattisgarh (3.2%). For snake bites, Maharashtra and Telangana (each 9%), Andhra Pradesh (8%), and Chhattisgarh (7%) had the highest percentages. Delhi (30%), Daman & Diu (21%), Lakshadweep (19%), and Andaman & Nicobar Islands (16%) had the highest rates of acute cardiac events in the country (**Figure 2B**).

### HMIS EM Population-Level Registration Rate

The national HMIS EM Population-Level Registration Rate was 8,935.66 registrations per 100,000 population, with Delhi recording the highest rate at 48,749.43 and Bihar the lowest at 2,058.92. North India had a wide range of EMD Population-Level Registration Rates, with Delhi having the highest. Goa was an exception in West India, with a higher rate than Gujarat and Maharashtra. Kerala in the south had a significantly higher rate than Telangana, which had a lower rate than its neighbors. The East and Northeast regions showed the greatest variation, with Arunachal Pradesh recording the highest rate and Bihar the lowest (**Figure 3A**).

**Figure 3:**
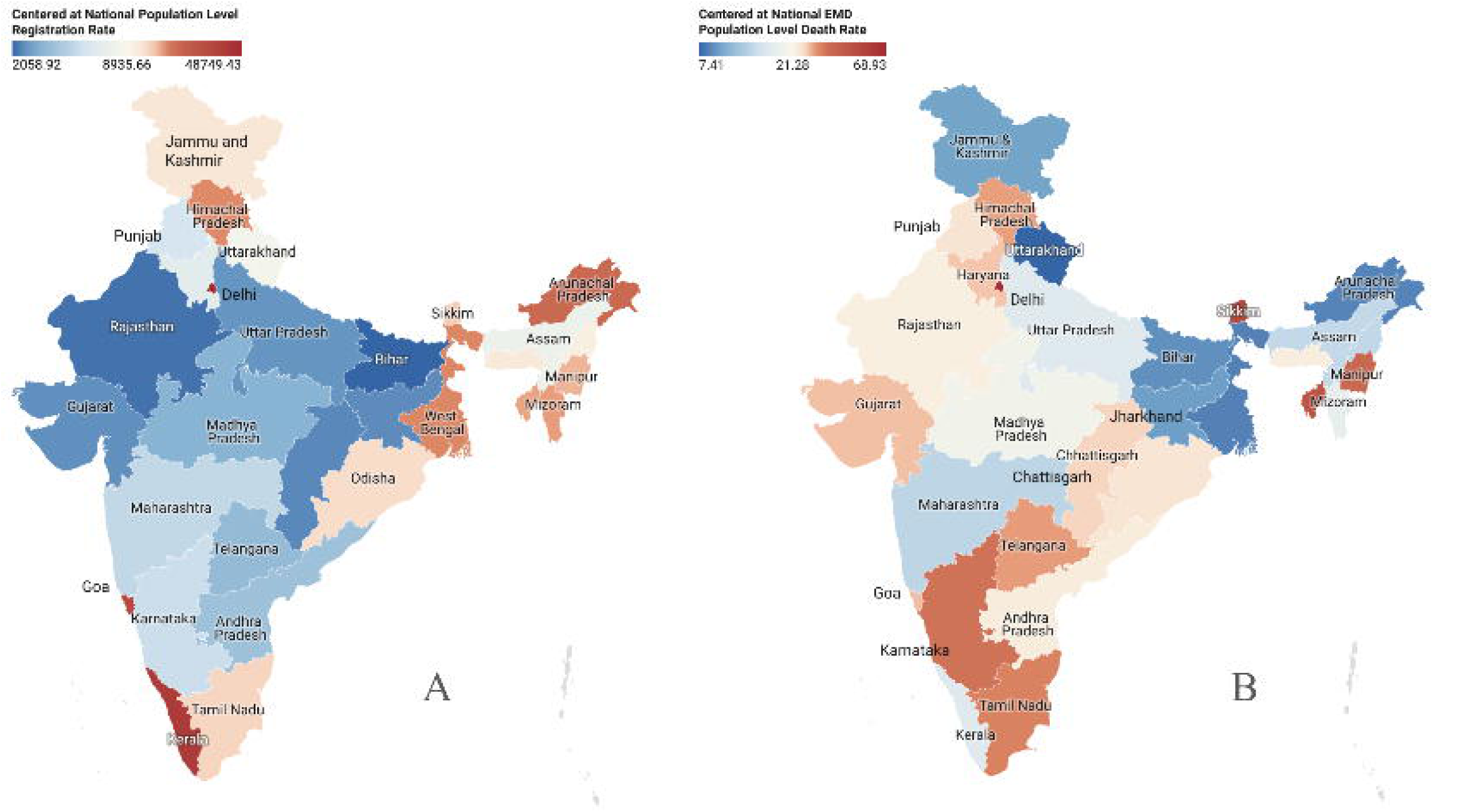
HMIS Emergency Medicine Department Population Level Rates for 35 States and Union Territories (2019-20), Including A) Registration Rates and B) Death Rates.

### HMIS EM Population-Level Death Rate

The national HMIS EM population-level death rate was 21.29 deaths per 100,000 population, with Delhi having the highest rate (68.93) and Uttarakhand having the lowest (7.41). Delhi had the highest death rate in North India. Goa and Gujarat in the west had higher death rates, while Maharashtra had lower rates. Karnataka had the highest death rate in the south, while its immediate neighbor, Kerala, had much lower rates. The East and Northeast regions showed the greatest variation, with Sikkim having the highest rate and West Bengal the lowest (**Figure 3B**).

#### GBD Descriptive Analysis

The GBD data reported 2,047,175,737 EMC incident cases in 2019, with 4,846,724 deaths. At the national level, EMCs accounted for 27.22% of all-cause disease incidence and 51.72% of all-cause mortality.

### GBD EMC Incidence Rates

Nationally, among the 29 conditions in this study, diarrheal diseases had the highest EMC incidence rate (120,789.43 cases per 100,000), while rabies had the lowest (0.40 per 100,000) (**Figure 4**). Regionally, Odisha reported the highest EMC incidence rate at 173,414.91 cases per 100,000, while Nagaland reported the lowest at 104,407.17 cases per 100,000 (**Figure 5A**). Thirteen states, particularly those in central and northern India, had EMC incident rates exceeding the national rate of 147,203.96 cases per 100,000. Seventeen states, predominantly in the South and in parts of the Northeast, had lower EMC incident rates than the national rate.

**Figure 4:**
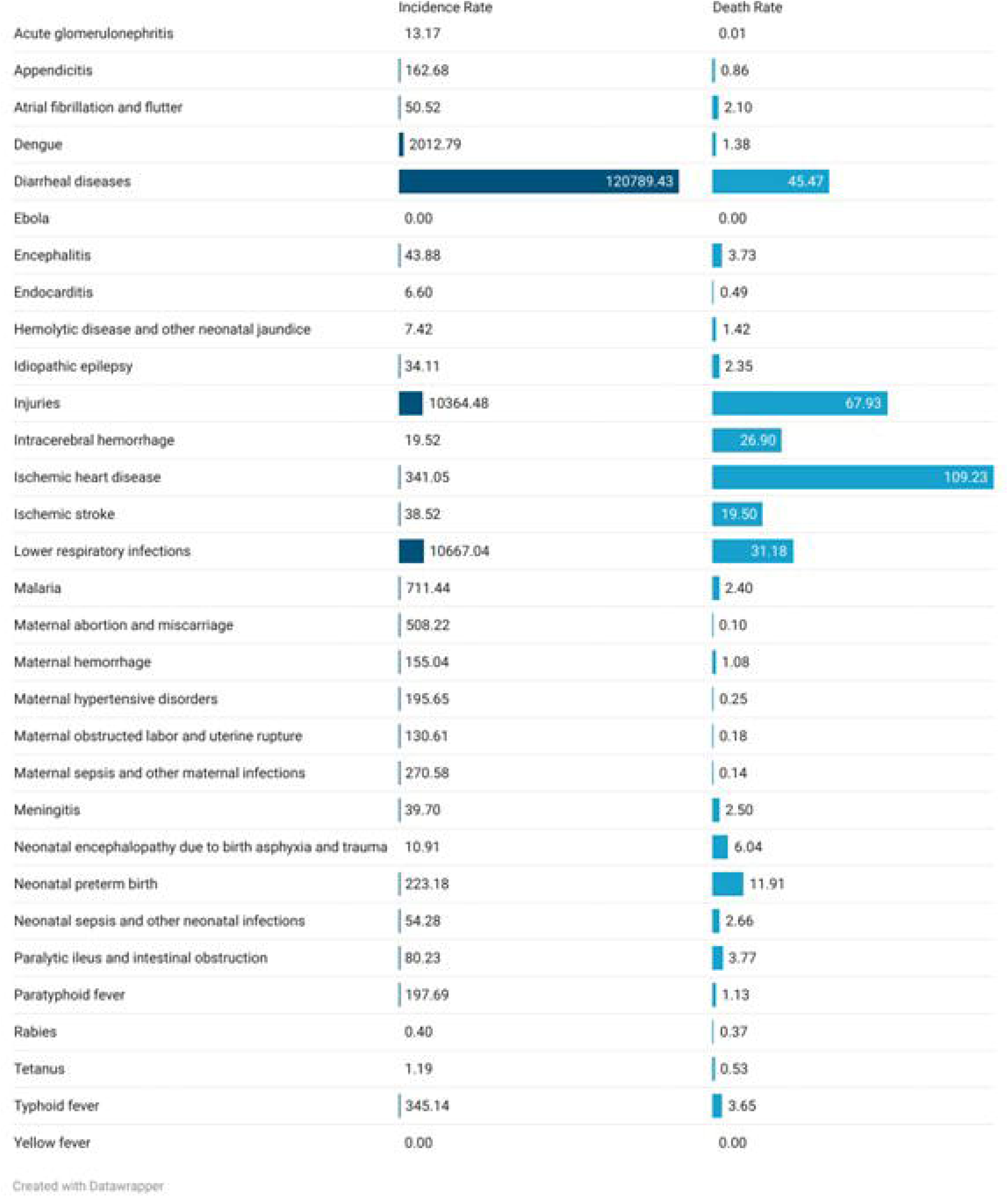
GBD Emergency Medical Conditions national rates (2019-20), including Incidence and Death Rates.

**Figure 5:**
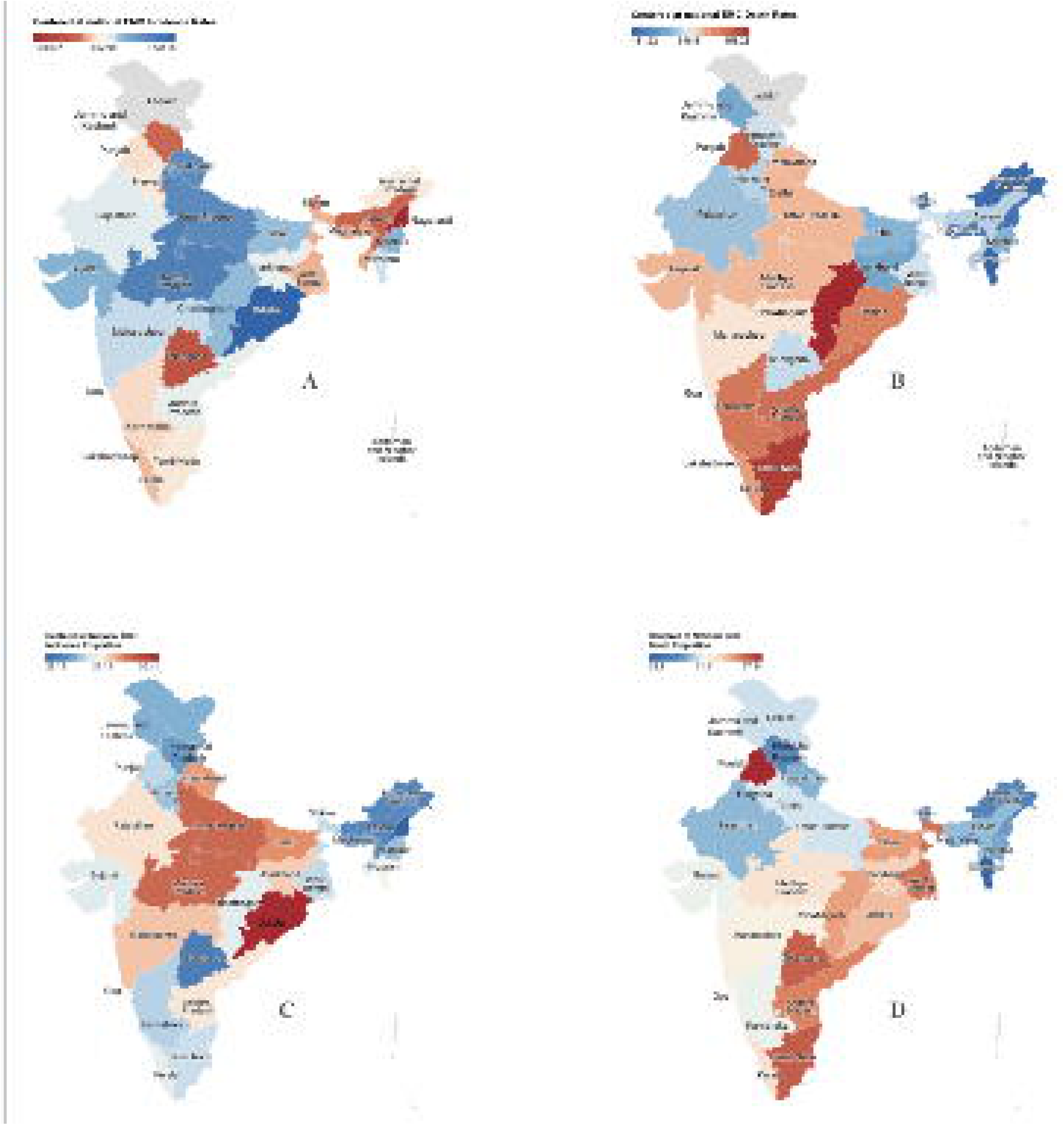
GBD Emergency Medical Conditions for 35 States and Union Territories (2019-20), including A) Incidence Rates, B) Death Rates, C) Incidence Proportion, and D) Death Proportion.

### GBD EMC Death Rates

Nationally, out of the 29 conditions for this study, ischemic heart disease had the highest EMC death rate at 109.23 deaths per 100,000, while Acute Glomerulonephritis had the lowest rate at 0.01 deaths per 100,000 (**Figure 4**). Regionally, Chhattisgarh reported the highest EMC death rate at 453.23 deaths per 100,000, and Mizoram recorded the lowest rate at 181.22 per 100,000 (**Figure 5B**). In total, 12 states reported EMC death rates higher than the national value of 348.51 per 100,000. Southern and Northeastern India, which had lower EMC incidence rates, showed higher death rates. However, Telangana in South India had lower incidence and death rates. Manipur and Arunachal Pradesh in Northeast India had high EMC incidence rates and lower death rates. Central and parts of North India showed higher EMC incidence and death rates than the national values.

### GBD EMC Incidence Proportions

The national GBD incidence proportion of EMCs was 27.22%. Odisha recorded the highest incidence proportion at 30.41%. Nine states had an incidence proportion higher than the national value. Nagaland had the lowest incidence proportion in the country (23.19%) (**Figure 5C**).

### GBD EMC Death Proportions

The national GBD death proportion of EMCs was 51.72%. Punjab (57.54%) and Tamil Nadu (55.96%) had the highest death proportion due to EMCs. Thirteen states surpassed the national death proportion. Mizoram (32.90%) reported the lowest death proportion in the country (**Figure 5D**).

#### Mapping of GBD EMC with HMIS EMD Registrations

In HMIS, trauma-related registrations comprised 8.22% of total EMD registrations, whereas injuries in GBD made up 7% of the total incidents. Obstetric registrations accounted for 1.86% of HMIS and 0.85% of GBD EMC incidence numbers. Emergency acute cardiac events accounted for 0.81% of HMIS EMD and 0.23% of GBD EMC incidence. Similarly, cerebrovascular accidents accounted for 0.56% of HMIS and 0.04% of GBD EMC incidence.

### Analyzing GBD EMC Incidence Proportion with HMIS EMD Registration Proportion

Sikkim, Andhra Pradesh, Uttarakhand, and Tripura had the highest difference between the HMIS EMD Registration Proportion and the GBD EMC incidence proportion. Nagaland, Punjab, Gujarat, and Assam had the least. Madhya Pradesh and Meghalaya reported higher HMIS EMD Registration Proportions than GBD EMC incidence proportions (**Figure 6A**).

**Figure 6:**
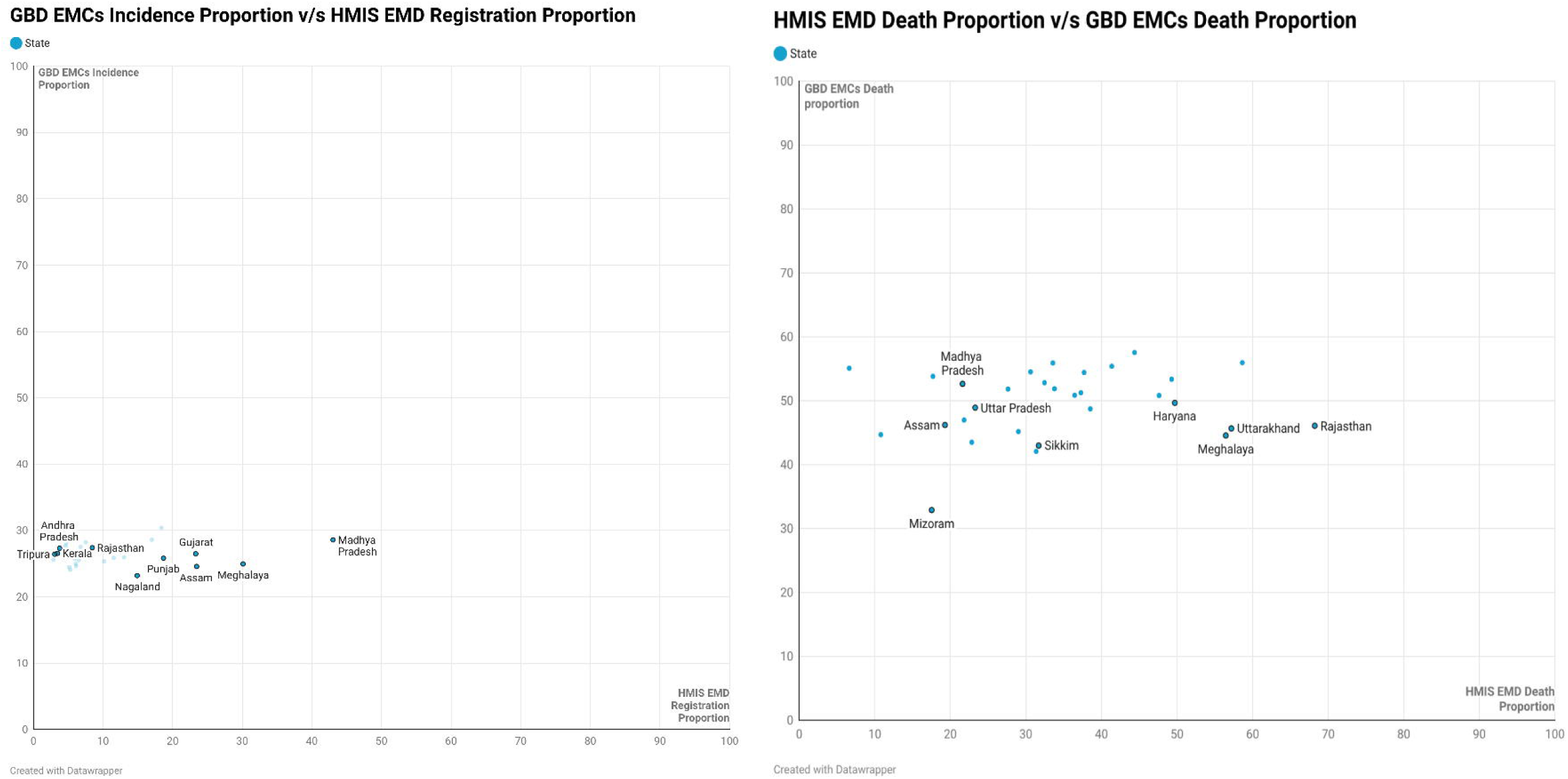
Comparison of data from HMIS emergency medicine departments and GBD emergency medical conditions for 35 states and UTs (2019-20) for A) Incidence Proportions vs Registration Proportions and B) Death Proportions

### Analyzing GBD EMC Death Proportion with HMIS EMD Death Proportion

Andhra Pradesh, Odisha, Nagaland, and Madhya Pradesh had the largest difference between the HMIS EMD Death Proportion and the GBD Death Proportion. Jammu & Kashmir, Jharkhand, and Gujarat had the least. Haryana, Tamil Nadu, Uttarakhand, Meghalaya, and Rajasthan had higher HMIS EMD Death Proportions than GBD EMC death proportions (**Figure 6B**).

## DISCUSSION

The primary aim of this study was to evaluate the national EMD service utilization in addressing the EMC disease burden, using two different data sources: GBD and HMIS in the 2019-2020 period, to investigate the correspondence between population-level and health system-level indicators for emergency medicine and the underlying data patterns and gaps. Analyzing the 2019-2020 financial year provides a reliable pre-pandemic baseline for understanding trends in both EMC disease burden and EMD service utilization. Our analysis reveals that only 119,103,358 HMIS EMD registrations were recorded, compared to 2,047,175,737 GBD EMC incident cases.

We observed variations between the two datasets, possibly for several reasons. Firstly, differences in numbers across sources could suggest inconsistent data collection and reporting practices, as well as on-the-ground differences in healthcare access and utilization. While the HMIS database is one of the few sources of information on health system functioning at national and sub-national levels, concerns have been raised about how HMIS data are captured. A 2013 policy brief highlighted critical issues affecting data collection in HMIS, including insufficient data from private hospitals and the submission of low-quality data.^9^ The poor data quality was partly attributed to inadequate training of ground-level hospital staff. In addition, HMIS reports often include limited data from private hospitals in India, which can affect estimates of EMC disease burden and EMD service utilization at the national and sub-national levels. The Uttar Pradesh (UP) state government’s district-level survey to identify on-the-ground barriers affecting HMIS data reporting by hospitals across the state discovered that during the 2013-14 period, no fixed deadlines were set for hospitals statewide to submit their monthly data. As a result, only 75 private hospitals regularly submitted monthly data reports to the government. However, UP’s Technical Support Unit introduced data collection training programs and established fixed deadlines for monthly report submissions, among several other measures, leading to nearly 2514 private hospitals in the state regularly submitting their data by 2018-19.^10^ Therefore, missing data and actual reporting errors in HMIS can possibly contribute to discrepancies in numbers between HMIS and other sources, such as the GBD.

Another potential reason for differences between sources could be barriers to accessing emergency care. Patients may be unable to access timely EMD services due to the “three delays” model: delay in deciding to seek care, delay in reaching an adequately equipped health facility, and delay in receiving care at the health facility.^11^ In 2022, Ravindra et al. reported an underutilization of pre-emergency and emergency services in India.^12^ This was attributed to a lack of awareness of ambulance availability and distrust in the reliability and care provided during transfers. HMIS can only record data for those who can access healthcare services. In settings such as India, where EMD service utilization is suboptimal, health information system numbers would not match and, more often than not, would undercount population-level case incidence.

Another reason likely stems from differences in how EMCs are classified in India (HMIS) versus in global data sources such as the GBD. While we used the standard framework to select 29 EMCs from the GBD and HMIS databases for this analysis, it is not grounded in local Indian EMC epidemiology or policy prioritization data.^13^ This can lead to classification differences, likely causing the exclusion of certain EMCs that may be of unique importance to India in our analysis. Additionally, EMC reporting can vary across hospitals and states because local clinical practice may define an “emergency” differently. This could subsequently impact HMIS numbers. The resolution of the data used in the current analysis precludes us from commenting on which of the above-mentioned issues (data reporting errors, limited healthcare utilization, and definition differences) are at play, or the extent to which each contributes to differences between HMIS and GBD for comparable outcomes.

### Policy Implications

Our findings underscore the need to strengthen EMD-level data collection to enhance emergency medicine service delivery at both national and state levels. Differences between EMD registrations and EMC incidence reflect utilization and/or reporting gaps. Implementing well-defined nomenclature and standardizing conditions can improve registration data quality by strengthening data infrastructure and training, and by ensuring that emergency protocols and treatment methodologies align with proposed national guidelines.^14^

High EMD death rates highlight the need for a focused strategy to improve EMD service utilization. National and state governments could invest in developing emergency medical service frameworks that outline standardized protocols for identifying, registering, and treating EMCs. A novel example is the Emergency Care Data Set (ECDS), implemented in the National Health Service (NHS) in England, which serves as a standard for Emergency Care.^15^ A reliable data registration, capacity evaluation, and EMC reporting verification process must be established at the hospital EMDs, district, and state levels. ^16^

State-wise variations in HMIS EMD registration and death rates reflect differences in health system capacity, accessibility, and population distribution. Higher EMD registration rates in Kerala, Goa, and Delhi reflect better per-capita EMD infrastructure and hospital availability. In comparison, lower rates in Bihar, Rajasthan, and the Northeastern states reflect under-resourced facilities and limited geographic access. Total population, public versus private utilization, and socioeconomic and cultural factors further influence service utilization.^17^

States with large gaps between EMD registrations (from HMIS) and GBD EMC incidence (from GBD), including Manipur, Goa, Arunachal Pradesh, Rajasthan, and Chhattisgarh, require tailored training programs and capacity-building initiatives to improve data reporting accuracy and close these gaps. Furthermore, states with high EMD death rates, like Haryana, Punjab, and Sikkim, require targeted interventions to reduce mortality rates by improving emergency care quality. On the other hand, states such as Meghalaya, Rajasthan, and Uttarakhand, which show differences between HMIS and GBD data, require further assessment to verify data accuracy.

### Strengths and limitations

In this novel nationwide assessment of emergency capacity and burden, we comprehensively analyze HMIS and GBD data and investigate their relationships with overlapping health indicators. The findings have direct policy implications, guiding strategic focus on regions that need to strengthen emergency departments and improve reporting mechanisms. This study also addresses the shortcomings in HMIS data and the need for standardization and improved reporting of emergency department conditions to streamline the allocation of emergency care resources.

While HMIS is based on health system data and relies on data reported by healthcare facilities, which vary in quality and completeness, GBD offers population-level estimates. Since India lacks widespread community-level reporting for EMCs, GBD data helps bridge this gap. Using data from both HMIS and GBD provides a more comprehensive understanding of EMCs in India while recognizing the strengths and limitations of each data source.

The study has several limitations. Firstly, the data used in this study are from 2019-2020. Only 9 of 29 GBD conditions were mapped to the 6 available HMIS conditions, limiting the study’s ability to accurately capture and represent the true numbers. This discrepancy may partly reflect GBD not accounting for India-specific EMC classification. Together, these factors further limit the comparability and reliability of the findings because underrepresented or missing data skew the overall assessment. The study also has discrepancies in the registration of certain conditions across GBD and HMIS, indicating gaps in data completeness and coverage. Moreover, HMIS data skew toward public-sector hospitals and do not fully capture private hospitals. Secondly, the absence of data from India’s Union Territories in the GBD database is a significant limitation, hindering a comprehensive understanding of national-level health trends and burdens. Lastly, we cannot assess the proportion of the disparity attributable to underreporting of the condition or to underutilization of healthcare services.

## CONCLUSION

EMCs account for 27.22% of all-cause disease incidence and 51.72% of all-cause mortality at the national level in India. EMC-specific data revealed significant disparities across GBD and HMIS databases. Establishing robust data registration processes, incentivizing accurate reporting, and strategically allocating resources are critical steps toward improving emergency care delivery.

### Relevance to Preventive Medicine

Our study highlights the need to establish a dedicated emergency medicine database in India to collect and store data from EMDs in public and private hospitals. Standardized data collection and verification protocols ensure high-quality data informs policies and programs that direct resources to strengthen infrastructure and capacity-building efforts, thereby ensuring equitable and timely access to emergency care services for patients across the country.

### Implications for Clinical Practice

Our study highlights variations in EMC disease burden and EMD service utilization across states, underscoring the need for local benchmarks for clinical practice. Further, the gap between EMC disease burden and EMD service utilization in any given state is the first step towards capturing unmet clinical care needs. Future studies with more localized data can further this line of work.

## Data Availability

All data produced in the present study are available upon reasonable request to the authors.

## Author contributions

SZ and JRNV conceived the study. SZ supervised the study. GU, PKK, TG, PN, and SZ extracted and compiled the data. TG, PN, GU, and PKK analyzed the data. SZ, GU, PKK, and AA drafted the manuscript, and all authors contributed substantially to its revision. SZ takes responsibility for the paper as the guarantor.

## Data Sharing Statement

The dataset, data dictionaries, and data analysis for this paper are available upon request by contacting Siddhesh Zadey, BS-MS, MScGH, at.

## Declaration on use of AI

The authors used Claude Cowork Opu 5 for data validation. No generative AI/AI assisted technologies were used for the writing process.

## Patient Consent

Not applicable

## Ethical Clearance Statement

No ethics approval was considered necessary since we used publicly available aggregate-level data.

---

**Table 1:** List of HMIS and GBD Outcomes.

| Source | Outcome | Formula |
| --- | --- | --- |
| **HMIS** | EMD Registration Rate | $\frac{\text{Number of Emergency Department Registrations}}{\text{Total Hospital Admissions}} \times 100,000$ |
| | EMD Death Rate | $\frac{\text{Number of Deaths in the Emergency Department}}{\text{Total Hospital Deaths}} \times 100,000$ |
| | EMD Death Proportion | $\frac{\text{Number of Deaths in the Emergency Department}}{\text{Total Hospital Deaths}} \times 100$ |
|  | HMIS EMD Registration Cause-wise | The number of registrations for specific medical conditions |
| | Cause-Specific Registration Proportion | $\frac{\text{Number of Registrations for a Specific Cause}}{\text{Total Emergency Department Admissions}} \times 100$ |
| | EMD Population Level Registration Rate | $\frac{\text{Number of Emergency Department Registrations}}{\text{Population}} \times 100,000$ |
| | EMD Population Level Death Rate | $\frac{\text{Number of Deaths in the Emergency Department}}{\text{Population}} \times 100,000$ |
| **GBD** | EMC Death Rate | $\frac{\text{Deaths Due to Emergency Conditions}}{\text{Population}} \times 100,000$ |
| | EMC Incidence Rate | $\frac{\text{Incidence of Emergency Conditions}}{\text{Population}} \times 100,000$ |
| | EMC Death Proportion | $\frac{\text{Deaths Due to Emergency Conditions}}{\text{Deaths of all causes}} \times 100$ |
| | EMC Incidence<br>Proportion | $\frac{\text{Incidence of Emergency Conditions}}{\text{Incidence of all causes}} \times 100$ |

**Table 2:** Mapping of GBD emergency medical conditions against HMIS emergency medical department registrations.

| HMIS Registered Causes | Corresponding GBD EMCs |
| --- | --- |
| Trauma | Injuries |
| Burns |  |
| Snake Bite |  |
| Obstetric Complications | Maternal abortion and miscarriage |
|  | Maternal hemorrhage |
|  | Maternal hypertensive disorders |
|  | Maternal obstructed labor and uterine rupture |
|  | Maternal sepsis and other maternal infections |
| Emergency Acute Cardiac Events | Ischemic heart disease |
| Cerebrovascular Accidents | Hemorrhagic stroke |
|  | Ischemic stroke |

